# Construction of patient trajectories to model clinical trial outcomes: Application to Myasthenia Gravis

**DOI:** 10.1101/2025.04.11.25325663

**Authors:** Marc Garbey, Quentin Lesport, Henry J. Kaminski

## Abstract

Accurate prediction of patient outcomes in clinical trials is crucial for timely assessment of treatment efficacy. This study introduces a novel approach to predict patient response by constructing temporal trajectories from longitudinal clinical data. We aim to extrapolate these trajectories to forecast individual outcomes and identify when new patients align with established response patterns. Utilizing data from the MGTX trial of myasthenia gravis patients, we evaluate the predictability of these trajectories and discuss potential confounding factors. Furthermore, our analysis yields an automatic clustering of patients based on treatment success, revealing potential associations with age and smoking status.

## 1. Introduction & Goal

Clinical trials are fundamental to evaluating new therapeutic interventions, yet they face numerous challenges, including patient heterogeneity, inconsistent data acquisition, and limitations in sample size. These issues are particularly pronounced in rare diseases, where small patient populations and variable disease trajectories complicate robust outcome predictions. Additionally, the variation of pathophysiological mechanisms across subjects is often poorly defined or unknown. Mathematical modeling offers the potential to enhance clinical trial planning and more deeply understand trial results. The most advanced model of a human subject would be a digital twin. Originally conceived by NASA, digital twin technology has been broadly adopted across various industries and is now being applied to healthcare.[1-4] Digital twins have demonstrated value in augmenting control arms in common disorders, including oncology, Alzheimer’s disease, and multiple sclerosis trials by reducing sample size requirements through synthetic control cohorts.[5,6,7] In rare diseases, modeling would be particularly impactful to enable more efficient clinical trials by identifying early responders, predicting treatment efficacy, and optimizing trial design.

We propose a modeling framework aimed at enhancing the efficiency and predictive power of clinical trials, using myasthenia gravis (MG) as a representative example of a rare disease. [8] MG is an autoimmune neuromuscular disorder caused primarily by antibodies directed towards proteins on the post-synaptic surface of the neuromuscular junction. Symptoms range from disabling visual impairment to life-threatening respiratory failure. Despite recent advancements, there remains a significant unmet need in MG treatment due to poor adverse effect profiles of existing therapies, wide variation in patient response, and the fact that nearly a third of patients remain treatment resistant. To our advantage, MG serves as a good starting point for our work in that (1) The fundamental pathogenesis of MG is well understood. (2) There have been several rigorously performed clinical trials over the past decade with potential access to date to allow model formation. (3) The specificity of AChR antibody testing provides confidence in patient diagnosis, which is critical for analyzing electronic medical record data. However, predictive modeling in MG faces challenges, including disease rarity, fluctuating manifestation over time, and the yet to be defined variation of autoimmune pathology among subjects.

We developed a predictive model for patient outcomes based on the concept of patient trajectory, or a time-based path tracking key clinical metrics, such as improved quantitative myasthenia gravis (QMG) and MG-Activities of Daily Living (ADL) scores, reduced prednisone dosage, or achievement of minimal manifestations. [9] [10] [11] All these metrics are validated outcome measures and used to varying extents across clinical trials of MG. Constructing models for clinical trials presents several challenges: inconsistent timing of metric acquisition across patients, inherent uncertainty in neuromuscular disease scoring, and human factors affecting data accuracy. Given that MG trials typically enroll around one hundred and fifty, or fewer patients, predictive modeling must balance the need for sufficient data diversity while ensuring that patient populations remain comparable.

To develop and validate our approach, we utilize the randomized, single-blind, phase trial of prednisone plus thymectomy versus prednisone alone (MGTX) trial dataset, which demonstrated the added benefit of surgical removal of the thymus based on overall lower prednisone dose and QMG scores.[9] The three-year trial serves to exemplify the complexities of clinical trial design and optimization with respect to model building, which in requires integrating numerous inputs—age, gender, disease duration, medical history, and diagnostic studies—to generate comprehensive disease trajectories with an ultimate goal to construct digital twins of MG patients, leveraging data from multiple clinical trials and electronic health records to enhance patient care and treatment decisions.

This investigation presents an initial step toward constructing patient trajectory models, assessing their predictability, and exploring their potential for extrapolation in future clinical trials. We aim to provide insights into key control factors that impact modeling outcomes, laying the foundation for broader applications in clinical research beyond MG.

## 2. Methods

### Data Set

A complete de-identified clinical data set of the MGTX (NS 42685) was provided to the investigators by National Institutes of Neurological Disorders and Stroke) [5] : This 3 years trial investigated the effectiveness of thymectomy plus prednisone versus prednisone in patients with generalized, acetylcholine receptor antibody-positive MG between the ages of 18 and 65. Subjects doing poorly had escalation of prednisone, and for severe worsening, treated with rescue therapies of intravenous immunoglobulin (IVIg) or plasma exchange. After year 1, azathioprine therapy could be added for subjects with poor treatment response as judged by the site investigator. One hundred twenty-six subjects were enrolled and followed over a period of 36 months. The primary outcomes were the time-weighted averages of the QMG score and prednisone dose. The ADL and QMG scores were to be done from study baseline at month M0, M3, M4, M6, and then every 3 months to end of study.

### General Approach

We require a **1)** data structure of an object/process, and **2)** a means of propagating that data structure through time to generate virtual trajectories (how a patient is doing). For **(1)** we built a mathematical framework that defines the state variable and control variable of the patient trajectory. The state variable has patient outcome and clinical information along with patient history and comorbidities. The control variable is what the physician or a patient can impact, for example drug choice, drug dosage, surgery, diet and likely other factors. **(2)** is described as a mathematical operator Φ. We project the subject’s data from the clinical study into a mathematical object; by design a projection provides a partial view, not an absolute reproduction of the real clinical patient condition. The projection must fulfill our goal of outcome prediction, such as the improved QMG, MG-ADL, lowered dose of prednisone, or achievement of minimal manifestations, which are common trial metrics. We developed a Unified Patient Trajectory Clustering technique that leverages data from several clinical trials to (a) compute an index of predictability for patient symptom evolution, and (b) quantify the impact of factors such as smoking, age and BMI on drug dosing and patient outcomes. We will report on our experience in this manuscript on the MGTX well documented clinical trial case. The method is summarized in Figure 1.

**Figure 1.**
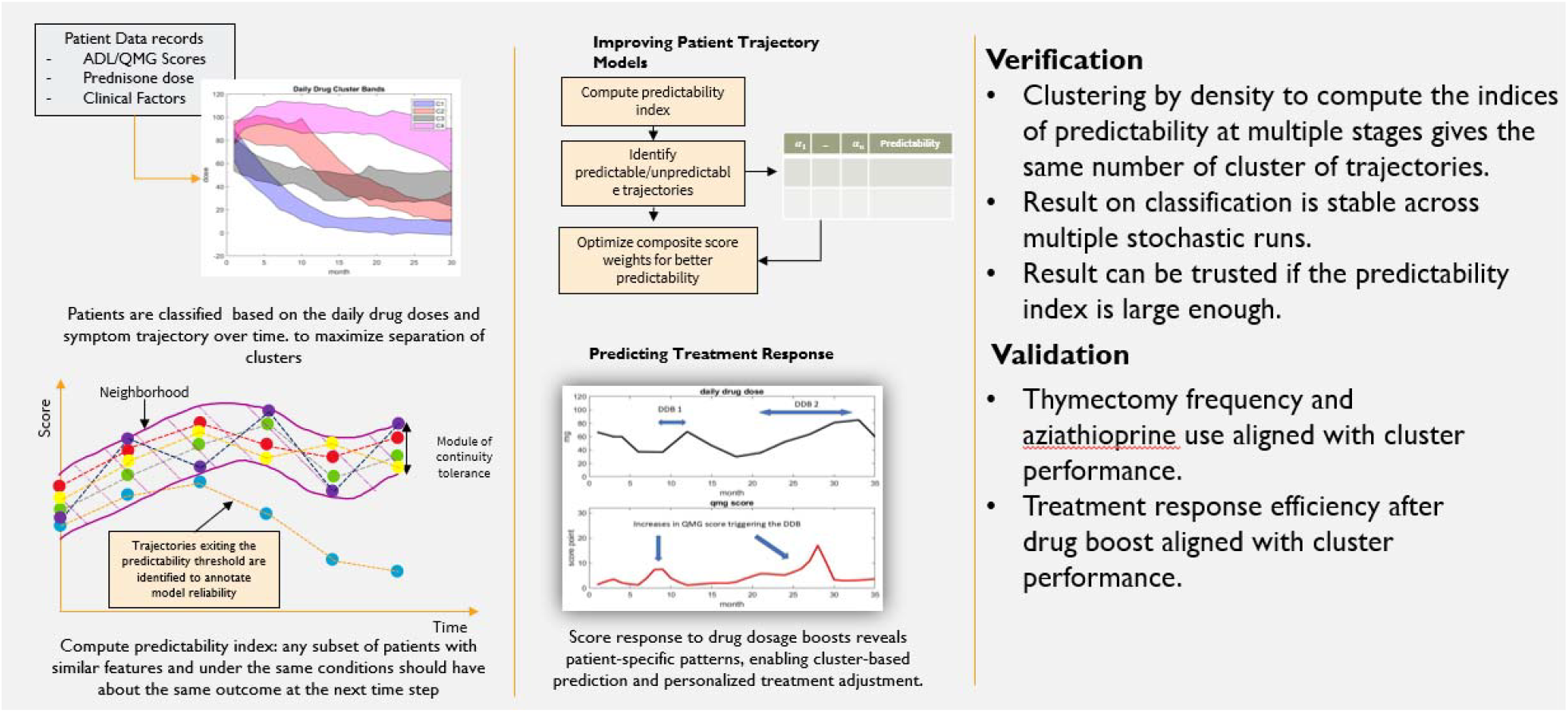
Model Development Process

### Patient Trajectory Technique

Let’s denote *V*_0_, *V*_1_, …., *V*_*q*_ the sequence of visits for a subject enrolled in a clinical trial, where q is the number of visits for data acquisition. Typically, the timing of visits varies to some extent for each subject.

The data set may require to go through a few steps of preprocessing: Trials often have missing entries, due to scheduling problems for example. The time interval between two visits of the same patient, could vary. We use standard interpolation techniques to project the state variable on a uniform time interval series and discard a subject data set when time gaps are too large to be filled with interpolation. The criterion is based on standard interpolation error estimates to stay within the observed noise level in the data set.

*Let* 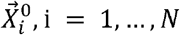 be the vector of features or state variable for patient i at initial time of the visit (*V*_0_).

*Let* 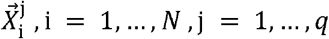 is the vector of features or state variable for patient i at time of visit *V*_*1*_ to, corresponding respectively to j = 1, .., q.

Thanks to the pre-processing step, the new vector feature is all set at the same time interval. Using the same notation 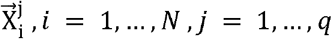, for this new vector feature, we can draw the patient trajectory *Pi*(*t*) as the set of state values progress with time.

A further step in preprocessing the data set might be denoising the time dependent trajectory with a standard filtering technique: we assume that *Pi*(*t*) is a continuous time dependent function in interval of time where no known singular events such as hospitalization occurred. Hospitalization may correspond to step value in the trajectory.

One can cluster the data set of trajectories *P*_*i*_(*t*) on a given time interval of continuous variation in order to check if patients may share similar behaviors: one starts with the simple K-means algorithm that aims to partition a priori the set of N trajectories into a given number Nc of clusters gathering patients that are the closest for a given norm. We will denote these Nc clusters 𝒞_*k*_, k=1,..,Nc, with all trajectories *P*_*i*_ belonging to one only .

We use the *L*_*p*_ norm, p = 1..∞ to evaluate the distance between two vectors: 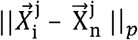.

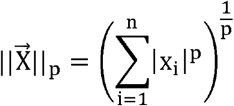

The quality of the clustering depends on the separation of the clusters: such indices of separation are the Davies-Boulin Index [12], the Calinsky-Harbasz index [13], or possibly the Dunn index [14]. All these indices use some variation of the silhouette function defined as follows [15]:

Assuming N clusters denoted 𝒞 _*k*_, k=1,..,N, for all trajectories *p*_*i*_ belonging to 𝒞 _*k*_, we set: 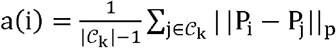, assuming the cluster is non-empty. | 𝒞_*k*_ | is the number of elements of the cluster.

a(i) represents the mean of the distance between *p*_*i*_ and the other trajectories within the cluster. We set:

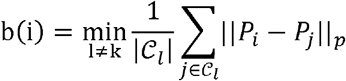

b(i) represents the mean of the distance of *p*_*i*_ to other clusters.

The silhouette value for each trajectory *p*_*i*_ is:

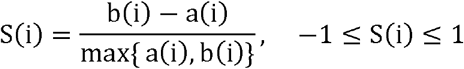

The best separation of clusters is characterized by how close to one are all silhouette values.

To optimize the clustering, we compute a quality index of the separation of the clusters and run the k-means algorithm within an optimization loop to maximize that index of separation. Trajectories for patients are small data sets, and one can run thousands of evaluations of the index of separation of clusters within minutes on a standard 11th Gen Intel(R) Core(TM) i7-1185G7 @ 3.00GHz processor.

Doing so we can address one of the deficiencies of the k-means algorithm, that is its sensitivity to the initial condition used in the iterative algorithm and as well to obtain the optimum maximum number of clusters.

In order to comply with patient safety, we decide to favor final clustering that may not have patient listed in the wrong cluster, which corresponds to negative values of the silhouette value S(i).

Rather than using the index of separation listed above we attempt to minimize our index defined as a weighted combination of patient signature: | | new*S* | |_2_, where:

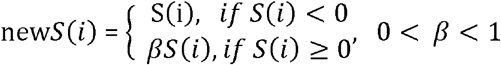

β is an arbitrary parameter to be chosen during the numerical experiment: the closer to zero the more we enforce the constraint on minimum negative silhouettes, the less we improve the separation of other patients. β and the number of clusters are decided with a trial-and-error method, as we like to minimize the number of patients with negative silhouette values as well. If we remove those patients from the model, it should not affect the statistical value of the conclusion.

Validation of the clustering technique is provided by visualizing the silhouette portrait as in [15] but also looking at the result that is the average trajectory for each cluster and its standard deviation across patient for that cluster. We need standard deviation along the trajectories that show clearly non overlapping average trajectories (in space-time).

### Patient Trajectory Predictability

We have assumed so far that patient trajectories can be clustered because they are not random. In other words, we have assumed that the patient outcome can be predicted along a trajectory. This is indeed not guaranteed. It would be mistaken to build a predictive model of the observation while the data set does not support any prediction.

A second part of our study is to compute a so-called index of the predictability of trajectory: If the data set supports predictability, the operator Φ exhibits some continuity property: any subset of patients with similar features and under the same conditions should have about the same outcome at the next time step; we can formulate that property mathematically: 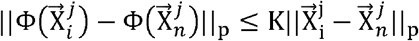 where K is the modulus of continuity.

We like to use K=1, to obtain a usable prediction to guess 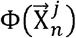 based on 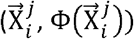 input.

First, rather than computing those distances on time at a given time step j, we should use block of time steps in order to limit the effect of local time fluctuation that is typical of the neuromuscular disease. In other words, we should look for a predictive model that is based on a few successive visits going from {*V*_0_,*V*_1,_ *V*_2_} for example to {*V*_4_, *V*_5_, *V*_6_}. It is straightforward to extend the *L*_*p*_ norm calculation in space and time.

Second instead of checking the continuity of the operator on single patients, which might be also too sensitive to noise in the data, we check that all sets of patients in the same neighborhood stay so at the time of the next interval of interest. The index of predictability will be defined as the percentage of patients or trajectories that stay so.

To implement this idea, we used a different type of clustering that paves the multi-dimensional space into a set of non-empty spheres of similar diameter. These spheres may overlap since we only want to test the continuity module of the operator. To compute a statistically significant module of continuity for the operator Φ, the partition of the space into spheres should maximize the density of trajectories that belong to each sphere. Eventually we can remove elements from each sphere to get a nonoverlapping partition of the data set of patients. Let us describe next the algorithm we have developed to show how we construct this partitioning.

Let us denote the partition of the data set 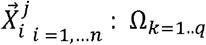 and let dist(Ω_*k*_) be the diameter of that subset. A meaningful partition consists of building subsets that have all similar diameters ≈ D and use proceeds based on density. For each trajectory *p_i_*, we compute the number of neighbor trajectories *p*_*l*_ that has the largest number of elements 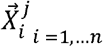. Once this sphere and all its corresponding trajectories are the exact same number of subsets for each time step. The iterative algorithm to generate that partition within distance D. One starts to build the partition with the sphere of diameter D centered on *p*_*i*_ that removed from the data set, we proceed computing the second most dense, populated sphere of diameter D, and so on, until we exhaust all possibilities. There are indeed multiple ways of partitioning the space into spheres. Our algorithm privileges density, in order to get the predictability index to be the most significant statistically.

We will discard our index calculation of all spheres that have a small number of elements, and therefore, do not support the verification of the continuity of the operator Φ. Similarly, D is a parameter that is chosen to get enough subjects per subset.

We can test if all mapped subsets 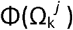 have a similar diameter at the next time window, which would correspond to a module of continuity of the operator Φ to be one. More precisely, since our algorithm is applied to a discrete data set, for each sphere of the initial partition, we check how many patient trajectories initially in Ω _*k*=1..*q*_ are outside the predicted sphere of center Φ (*p*_*i*_,) and diameter D at the next time interval. We define the percentage of the patient who stays within their sphere from one time window to another as the predictability index denoted Ip. A predictability index of 100% corresponds to the module of continuity of the operator Φ to be one. The smaller the predictability index Ip the less reliable is the extrapolation we can do based on mean trajectories.

### How to improve the trajectory model

This index of predictability can be used to select the best data set to build a predictive model. One can formulate this as an optimization problem that can be achieved by one of the standard stochastic optimization algorithms, such as the genetic algorithm [16], which is a method for finding solutions for constrained and unconstrained optimization problems that are based on natural selection. Alternatively, the data set of subjects can be partitioned into two categories: those in whom we can use extrapolation of trajectories and those that do not fit. From this partition, we can mine data on the control variable subset 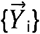 and corresponding patient data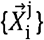 to extract some statistics of the features that may provide an explanation on the lack of predictability of these subject trajectories.

We may find that the combination of both techniques is best to optimize selection of variables that drive predictability of the model.

Another option we have tested is to focus on the prediction of a score and adjust the weight of each feature of that score to increase the quality of the prediction, i.e., maximize the predictability index. The ADL score, for example, is assembled from the individual score based on separated evaluation of muscle groups. The standard ADL score takes the sum of 8 such metrics, but there is no specific rational to say that the ptosis score should be as significant than the sit to stand score. With our methodology, we look for the optimum weight combination of these 8 individual scores that predict best the patient condition at the next time interval. Let α be the unknown 8 components vector and *ADL*_*j*_,*j* = 1,..,8 the individual score in [0,3] for each muscle group.

The optimization problem writes:

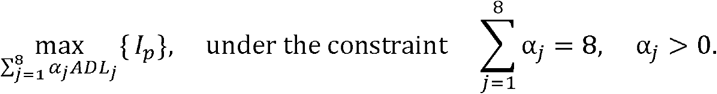

In our model, the weight vector should be independent of the patient and time of the visit indeed. We will define this modified ADL score: 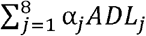 as a new ADL composite score. Following the same idea, we can combine the ADL score and QMG score into a weighted combination that maximizes the predictability index.Once again, the finding of the optimum weight factors can be formulated as an optimization problem:

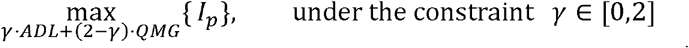

However, one technical difficulty related to the use of the control variable 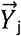. Subjects may vary in sensitivity to a treatment with the same exact condition 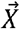. Some subjects might be highly responsive to a small drug dosage, while others require a larger dose. A possibility to get rid of that scaling factor would be to normalize 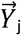 for each patient, within the same cluster, using the total amount of drug or thymectomy status. We emphasize this point as it is unquestionably the case for corticosteroid responsiveness and thymectomy response.

### Toward a model to generate patient trajectories

We will now introduce a new concept to construct a simplified, first-principles mathematical model to predict patient score trajectories from daily drug administration. This model is intended to serve as the foundation for a readily implementable tool that optimizes drug dosage according to patient-specific disease severity, thereby enhancing disease control.

The MGTX protocol for prednisone dosing was decided a priori by a consensus decision of experts and was as follows: Participants who were not already receiving prednisone at study entry received an alternate-day dose of oral prednisone starting at 10 mg and increased in 10-mg steps to 100 mg on alternate days or to 1.5 mg per kilogram of bodyweight, whichever was lower. For participants who were already taking prednisone, the dose could be increased up to 120mg in those who did not reach minimal-manifestation status by month 4. Minimal-manifestation status (MMS) is defined as “no symptoms or functional limitations from myasthenia gravis, but there may be some weakness on examination of some muscles” [17].

At month 4, the prednisone dose was maintained until minimal-manifestation status was reached and the QMG score was less than 14 and had also fallen at least 1 point below baseline, as determined by an evaluator who was blinded to the trial-group assignment. The alternate-day prednisone dose was then reduced by 10 mg every 2 weeks until a level of 40 mg was reached, with subsequent slowing of the taper to 5 mg every month, as long as MMS was maintained. If MMS was lost, the alternate-day prednisone dose was increased by 10 mg every 2 weeks until the status was restored. Tapering could resume 4 weeks later.

Once prednisone tapering commenced, the total dose of pyridostigmine, a cholinesterase inhibitor that produces rapid but transient improvement in strength, could not exceed 240 mg per day. Plasmapheresis or intravenous immune globulin was permitted at the discretion of the unblinded neurologist in patients whose condition was unstable, but it was not permitted to maintain MMS. Patients who did not have MMS at 12 months or who had an unacceptable level of side effects with prednisone could receive azathioprine at a dose of 2.5 mg per kilogram per day or another immunosuppressant such as cyclosporine if azathioprine caused unacceptable side effects.

To model patient trajectories, we must quantify the patient’s score response to drug dosage increases. The existing protocol employs a function Y(S,t), where S represents the score and Y the daily drug dose, which is set a priori the same for all patients. To develop patient-specific dosage regimens, rather than relying on generalized guidelines, we analyze the score response to isolated drug dosage boosts (DDBs). Ideally, the score should decrease following a DDB, though this is not always observed. Utilizing the MGTX dataset, we extract 78 DDBs and corresponding score responses to identify patterns in this relationship.

We hypothesize that these patterns are correlated with disease severity, specifically the patient clusters previously identified. Figure 2 illustrates the concept, showing daily drug dosage and score fluctuations for a patient in cluster 3. While a DDB should ideally be followed by a score decrease in responsive patients, clinical practice introduces variability, as clinicians adjust dosages based on experience to ensure patient safety. Nonetheless, we obtain multiple dynamic response samples from longitudinal datasets, which serve as the fundamental units for constructing cluster-specific response maps.

**Figure 2.**
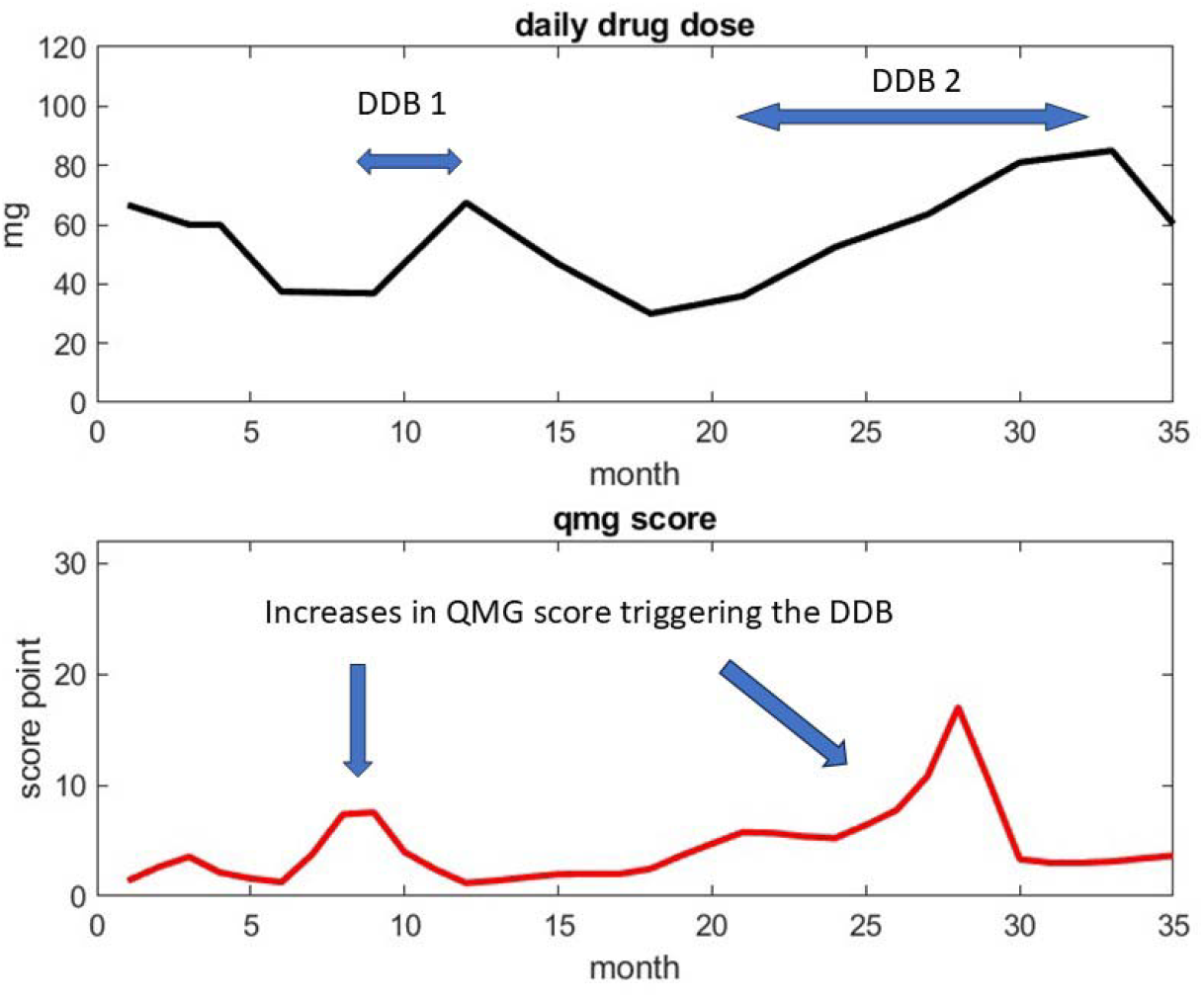
Example of Daily Drug Dosage and Drug Response with one patient from cluster 3. The first DDB occurs at month 10 as a result of the sudden increases of the qmg score at month 9. The second DDB occurs at month 21 and represents a continuous but moderate increase of daily drug dosage. One appreciates that the decision of the clinician on increasing drug dosage from QMG score observation reflects his own appreciation on risk of drug increases.

Given a sufficiently large database of dynamic responses per cluster, we can create a predictive map of drug dosage strategies that aim to control the score without relying on universal protocols. To assign a new patient to a cluster, we first compare their initial trajectory to reference trajectories (Figure 3). Subsequently, we verify that the patient’s DDB-to-score-change (ΔS) response patterns align with the cluster’s DDB database throughout treatment. This approach forms the basis for a novel machine learning algorithm for personalized drug delivery management.

**Figure 3.**
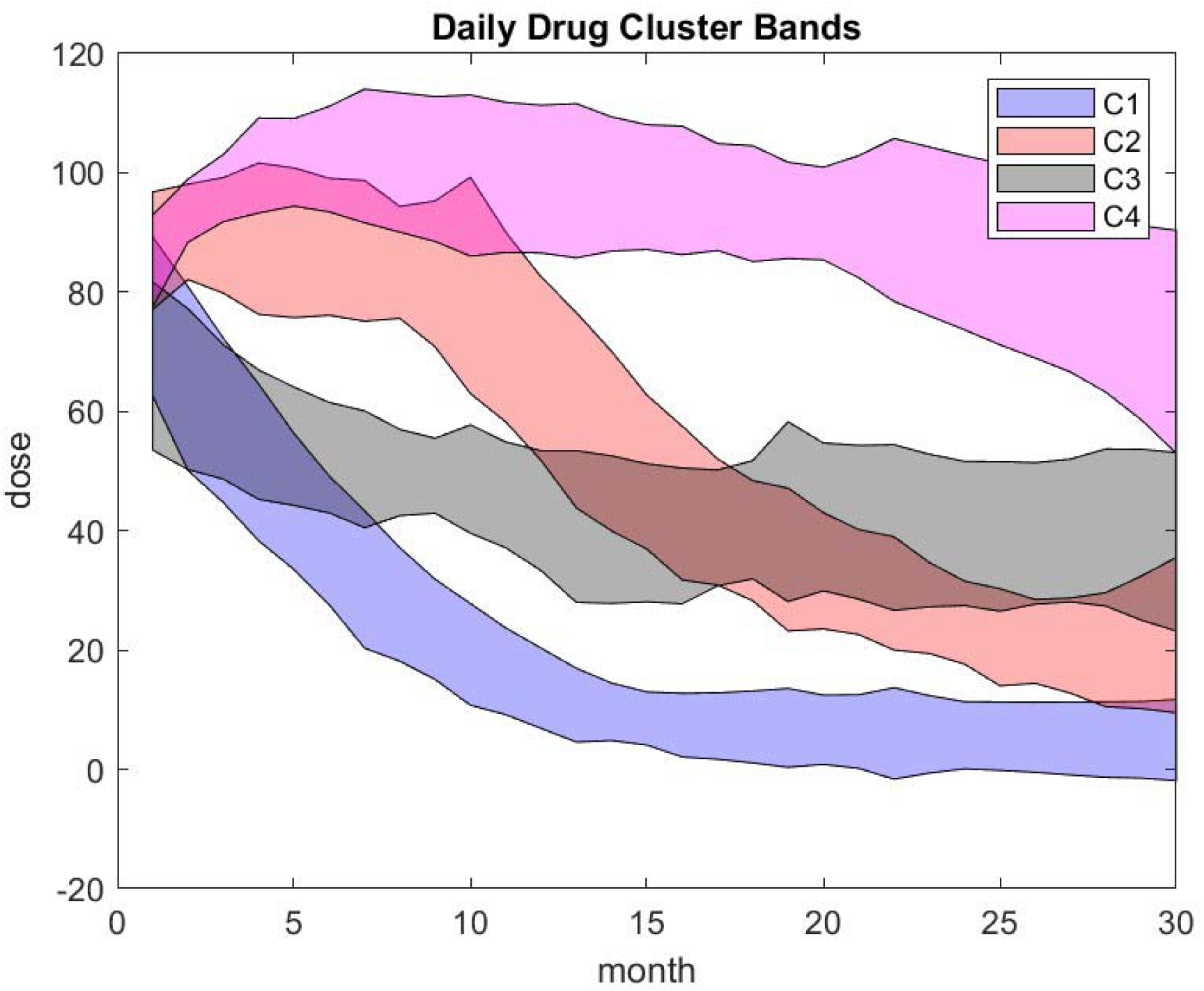
Patient trajectory over time representing the daily prednisone dose Corresponding to the 4-cluster result. Centroid of clusters represented by the solid curve corresponds to the mean patient trajectory of that cluster. Dotted line stands for the upper and lower trajectories estimate of the cluster within one standard deviation.

Next, we describe the main results obtained with our novel methodology.

## 3. Result

### Clustering Analysis Based on Daily Prednisone

We start the analysis by clustering the trajectories of the daily prednisone dose. The distribution of the MGTX patient population with respect to age and BMI is shown in Fig A1 of the appendix.Seventy-one percent of subjects were women, consistent with demographics of MG.

Out of 126 subjects, 111 patients have a record available covering 140 weeks. We exclude from the analysis the first 10 weeks, which was the time period of required dose adjustments, and the unlikelihood take an emerging pattern could be observed. K-means algorithm is run a thousand times and we chose the best weighted combination of patient signature corresponding to β = 0.1.The maximum number of clusters that give a good separation is 4: out of 111 patients 5 have negative silhouette function – see Figure 4.

**Figure 4.**
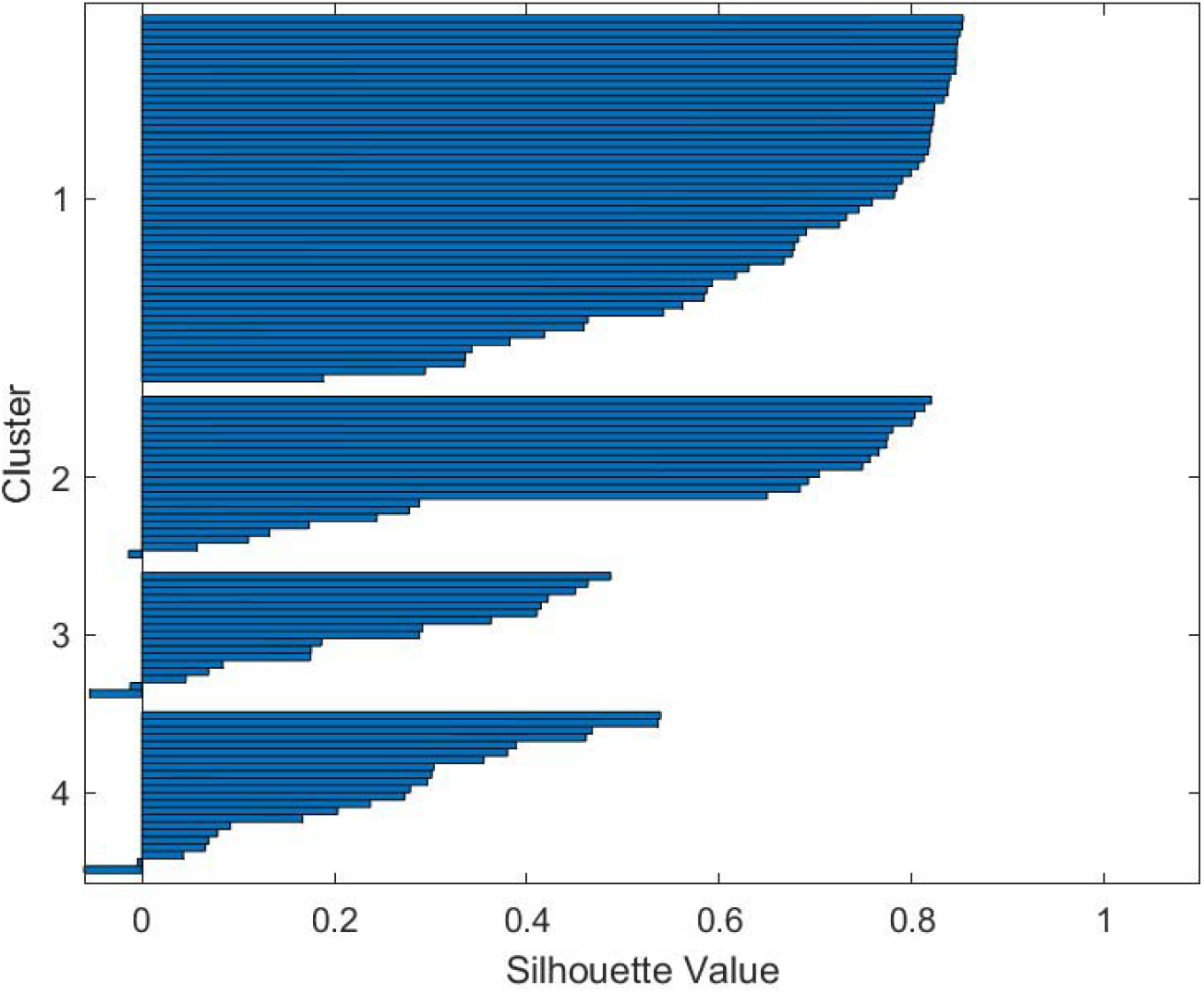
Silhouette value for the 4 clusters based on daily prednisone dose; The MGTX trajectory was computed with 111 patients that had complete record for the 36 months period. Five patients out of 111 have negative silhouette which means they are not properly classified. They are removed from the analysis of the impact of age, sex, smoking and BMI.

We compute the index of predictability on daily prednisone dose using a partition with subsets of diameter of the order of 8% of the maximum dose. This diameter guarantees that there is enough spheres with 10 patients or more to classify the trajectories. The algorithm shows that the number of spheres in the partitioning varies from 3 to 4 with the time interval of prediction, which is coherent with the k-means algorithm result.

The index of predictability for the primary outcome of MGTX is close to 90% and confirms the validity of using patient dose trajectory as a model.

The strip centered on the mean trajectory of bandwidth +/- one standard deviation for each 4 clusters is given in Figure 5. We order the cluster tags from 1 to 4, to correspond to the best to worst performance outcome. The numerical result of the clustering is stable, i.e. it gives the same computational result for each run. The sensitivity of the k-means algorithm on the initial guess had been overcome thanks to our method. We observe that no matter how small β is, there could be patients with negative signatures, but the impact of the 5 patients with negative signatures with the choice β = 0.1 on these curves is negligeable. Nevertheless, we removed these 5 patients from further analysis.

**Figure 5.**
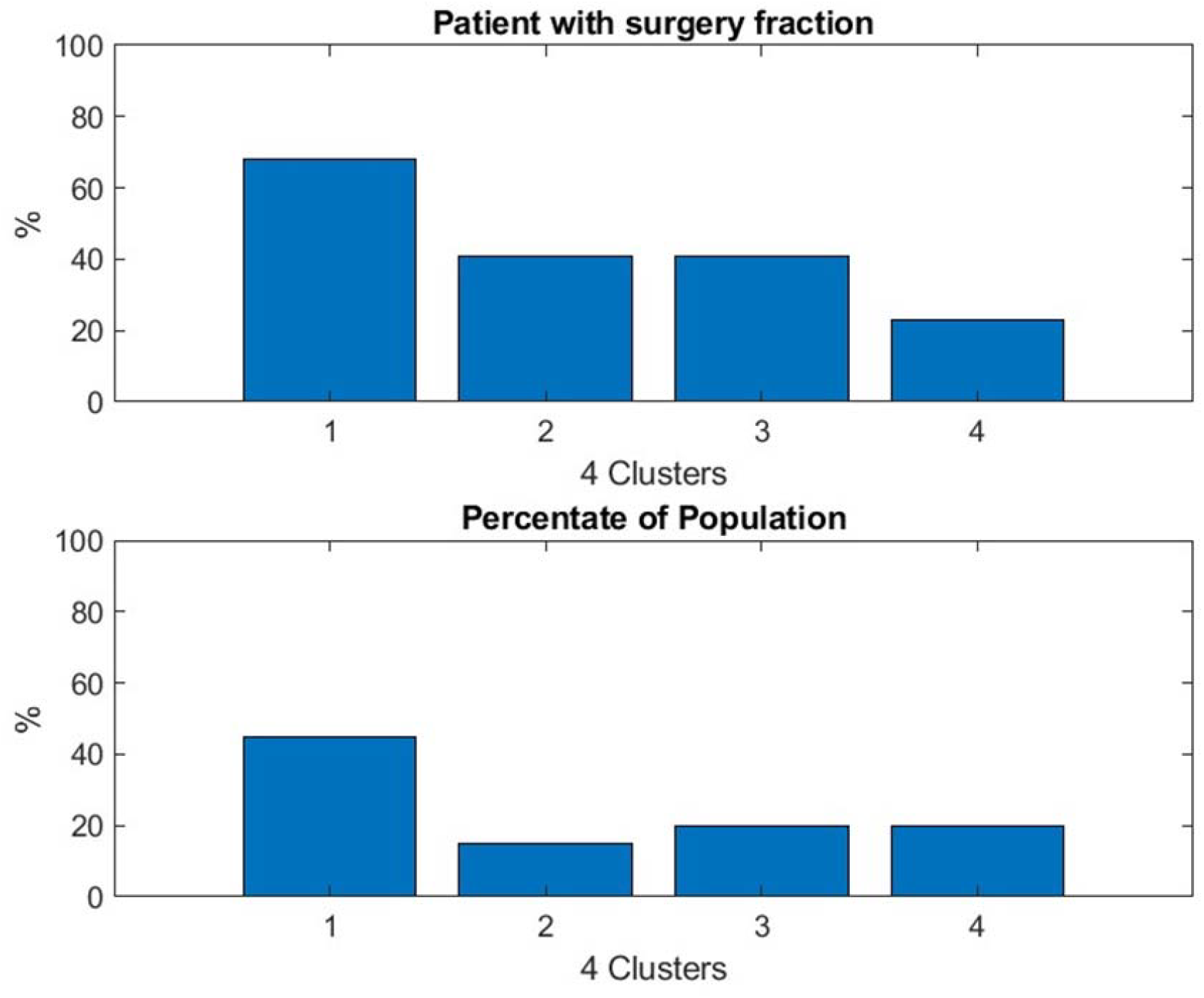
Result confirms the benefit of thymectomy for patient outcome expressed as daily prednisone dose. The thymectomy patients were more commonly in Cluster 1, which identified the subjects who responded best to treatment.

Almost half of the patients belong to cluster 𝒞_1_ and have the best daily prednisone dose outcome. The pattern of the other 3 cluster trajectories is clearly different and well differentiated. From this result we can search for the features that some of the patients in each cluster have the most in common.

First, we found that thymectomy is beneficial consistent with the primary outcome of the MGTX trial in [5]: the percentage of the patients who went through thymectomy decreases with the performance of the cluster – see Figure 5. As expected, the average number of doses of azathioprine, a metric of poor response to treatment, received by the patient increased as the cluster performance decreased - see Figure 6.

**Figure 6.**
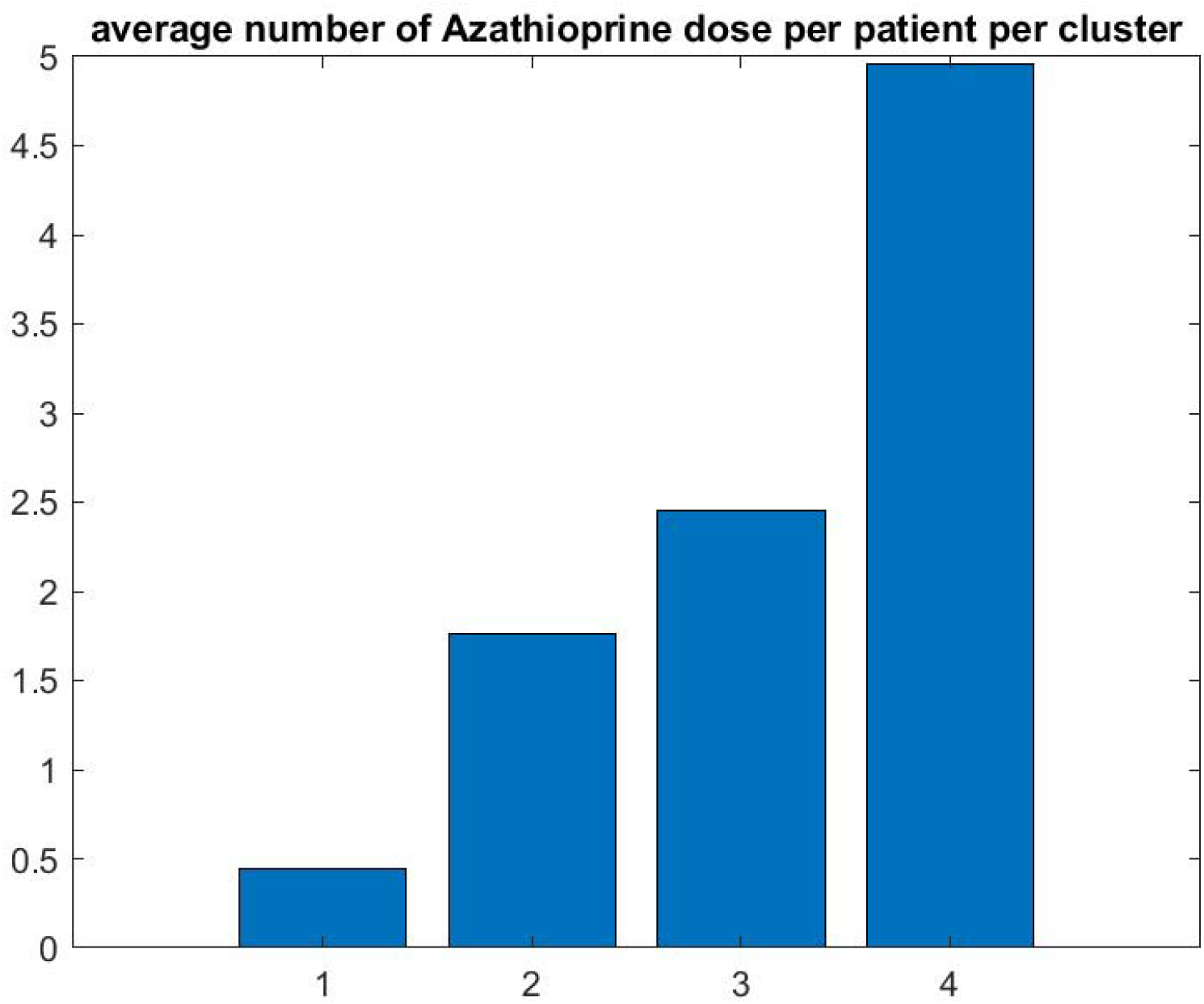
Graphic confirms that the patients who do best (respectively worse) have the lowest (respectively highest) number of doses of azathioprine

We asked whether smoking, BMI, sex and age play a role in cluster performance given their established risk factors for autoimmune diseases. We found that smoking had a moderate negative impact see Figure 7. The best cluster 𝒞_1_ has the largest proportion of patients below 50. Sex had little effect on the percentage of female per clusters with respectively 68%, 71%, 77% and 73% for cluster 1 to 4.

**Figure 7.**
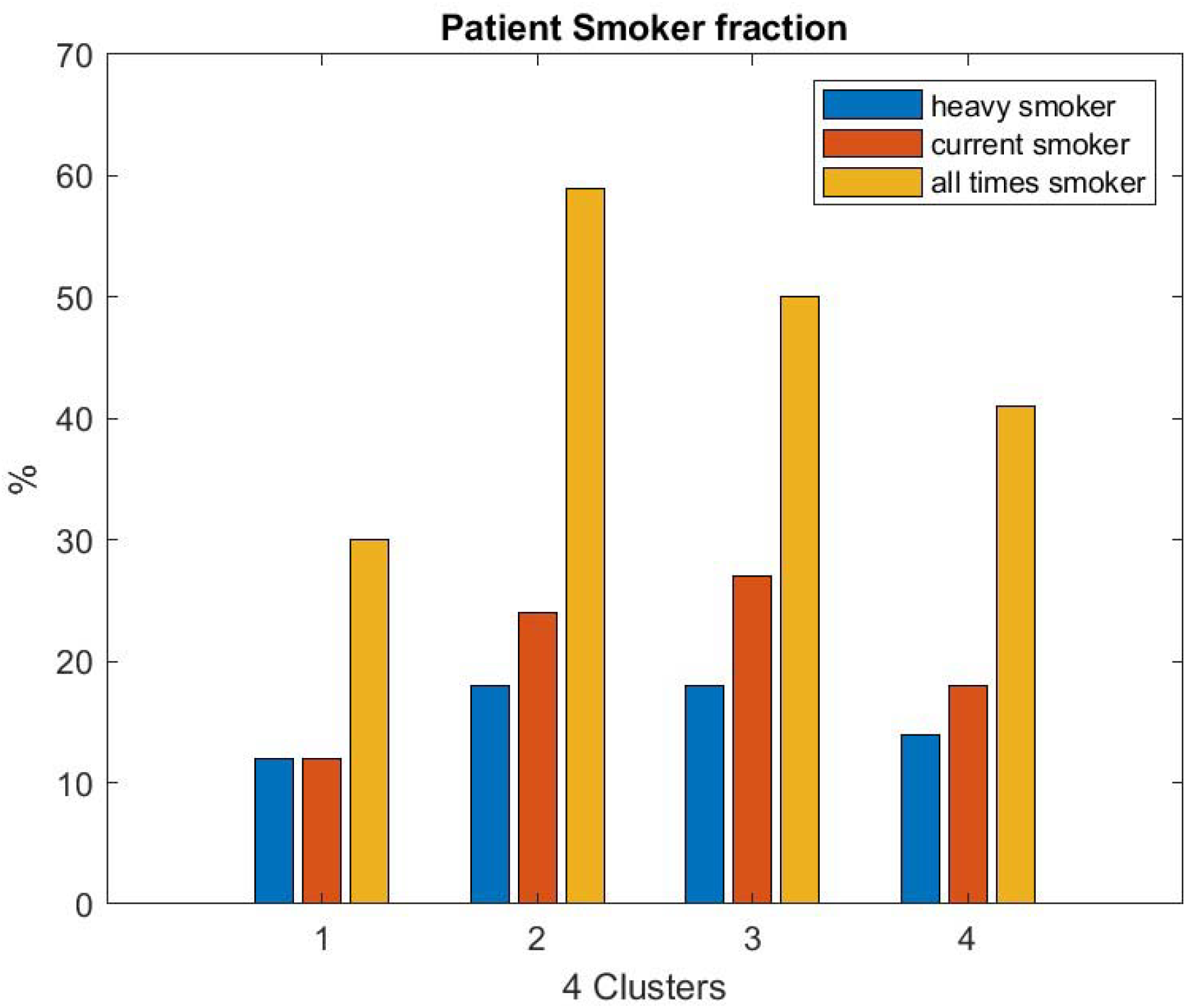
While this graphic shows some variation, half of the population of the clinical trial that belongs to the best cluster has the least number of smokers.

The average BMI of the population of each cluster seems to decrease somehow with the performance of the cluster. This cannot not be the indirect effect of a relationship between age and BMI: we check that the average BMI of the MGTX patient below 50 and above 50 it is not statistically different, but the distribution of these features is not a normal distribution as shown in Figure A1.

We found a temporal pattern of the dose of azathioprine during the MGTX trial: once a patient is on this drug, they typically maintained the drug. We can then define the time of failure of treatment as the time patient starts azathioprine. Figure A2 shows this pattern where each row represents a patient: patient have been ordered from late failure at the bottom of the left graph to early failure at the top. The 70 patients who never received the drug are not shown in the graph. The right graph plots the accumulated dose of prednisone received by patients at the time of receiving azathioprine. There appears to be a saturation effect on the accumulated dose of prednisone that corresponds to the time of failure of the treatment – see horizontal red line in Figure A2 right.

### Clustering based on MG-ADL or QMG scores

The ADL has eight individual categories to its score while the QMG has thirteen. We found initially that the clustering on individual ADL or QMG score items, such as eye lid drop, double vision, or arm weakness usually failed with either the clusters not being well separated or having a chaotic temporal behavior. Accordingly, the index of predictability of the ADL score is low and rarely reached 50% overtime. By optimizing the weighted combination of ADL individual test scores, we were able to improve that index of predictability. The optimum weighted composition (see Figure A3) provided a 70% average performance measured with the *L*_2_ norm. It should be noted that we did a relatively coarse search on the optimum weight. The problem has 8 unknowns and one constraint: the overall sum of coefficients stay fixed. Nevertheless, the clustering of trajectory for the new weighted ADL score gives satisfactory results with three clusters – see Figure 8. The number of patients with negative silhouette function is 11 out of 111. As we look at the properties of population feature in each cluster, we found that the impact of thymectomy on ADL score and the need for azathioprine is about the same as previously determined using the daily drug dose. The ADL patient score, that is the subjective report of the patient over the previous 14 days, is a much noisier signal than the daily prednisone dose that follows a protocol based on the QMG score corresponding to the clinical examination.

**Figure 8.**
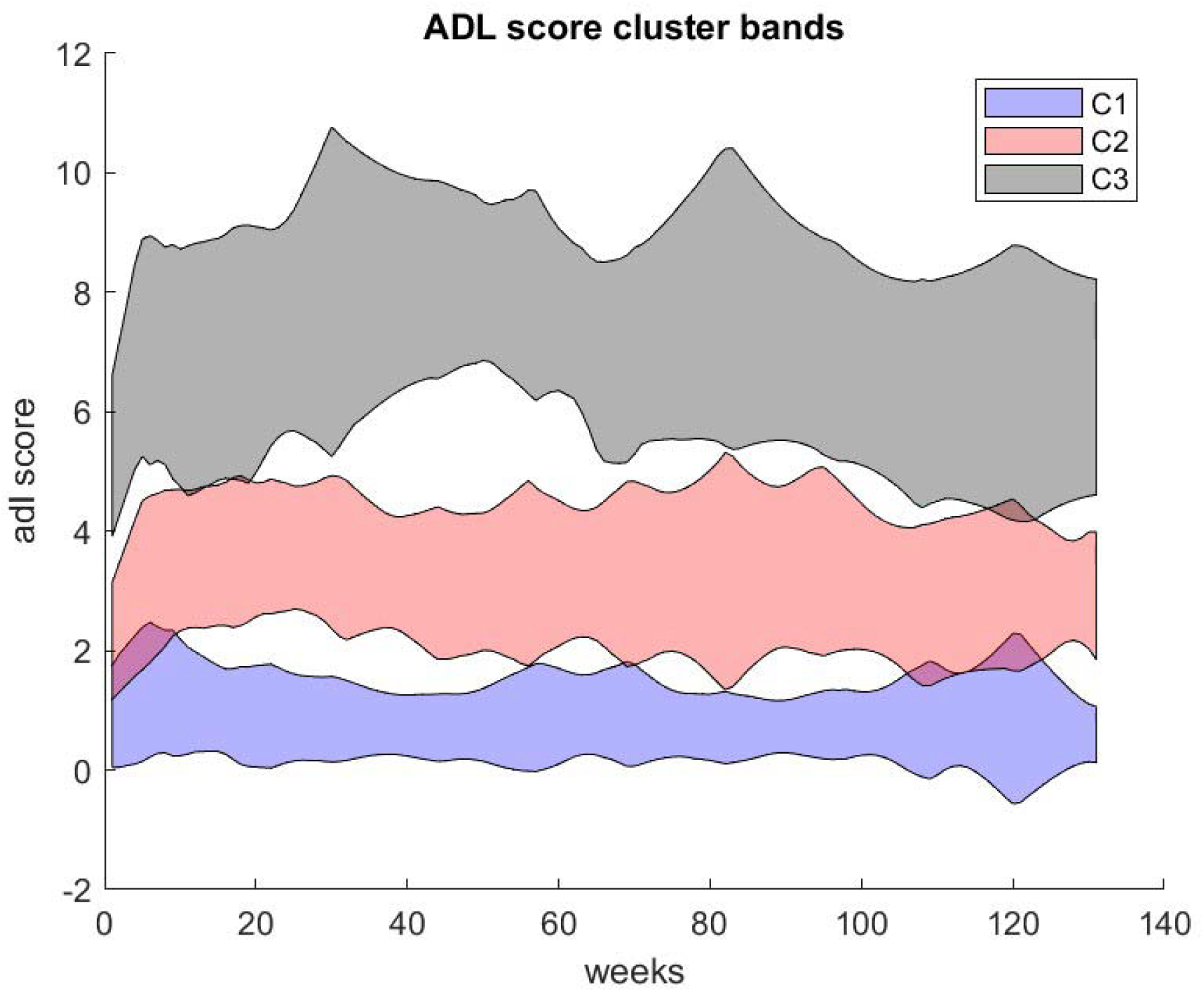
Composite ADL (secondary outcome of MGTX) score clusters with nice separation showing a moderate relative variation over time.

One can check on how this may translate in patient trajectory of daily drug dose versus ADL score using the same number of clusters for each metric. Since the new ADL score can be well described with 3 clusters, we use the same number of clusters for the daily drug dosage. The separation with three clusters for the drug dosage trajectories is even better than with four as expected. The heat map of Figure 9, gives the distribution of patients at the intersections of the daily drug clusters and score clusters with QMG – left panel, respectively ADL – right panel. This diagram sums up the results for 95 patients out of 111, as we want to keep only the patient with positive silhouette function for both metrics. As expected, the number of patients that belongs to the best cluster for both daily drug and composite ADL outcomes is the largest. However, even for the best drug trajectory cluster, see first raw of the heat map (Figure 9)- one can appreciate that some patients’ ADL scores are poorly controlled. We suspected that these patients would have benefited from a larger daily dose of prednisone or additional therapy. All patients outside the diagonal, would correspond to a suboptimal control of the drug dosage that was defined a priori for all subjects in the MGTX protocol. We obtain similar results using the QMG score.

**Figure 9.**
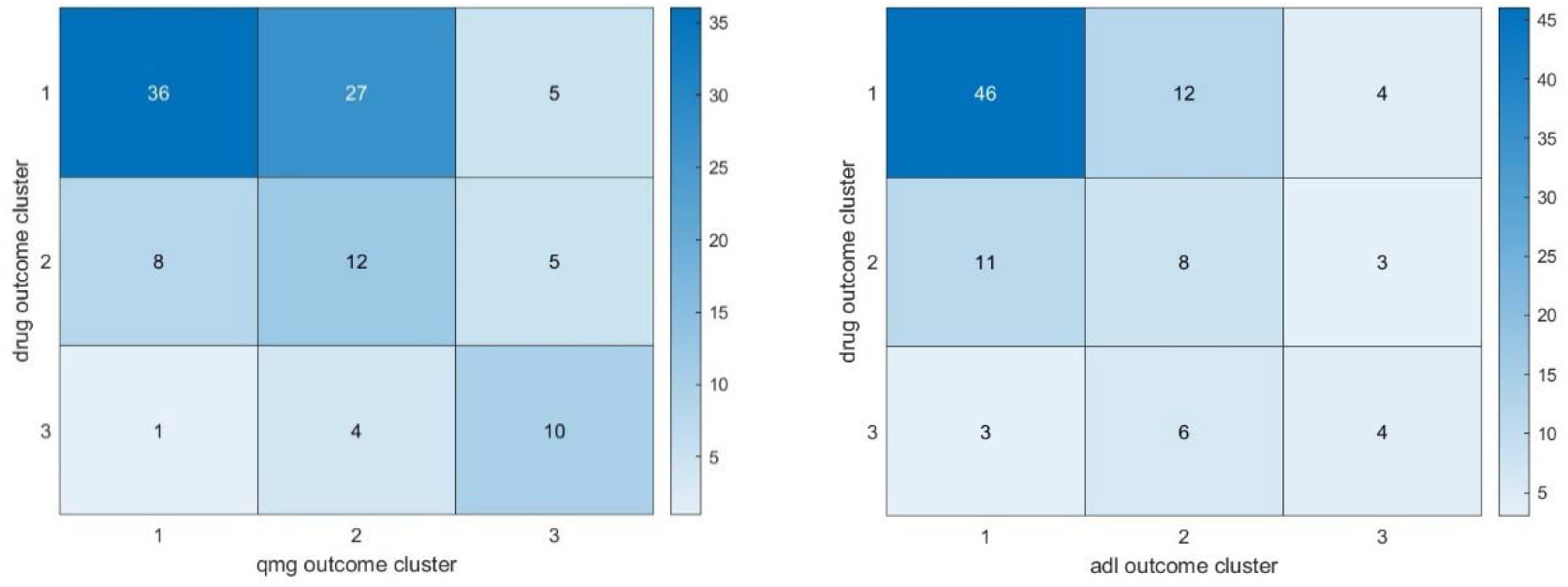
To compare the MGTX patient distribution between clusters for the daily prednisone dose, and the the adl-composite score, we use a 3 clusters algorithm for both outcomes. This diagram sums up the results for 97/108 patients. 5 patients (respectively 11) had negative signature in the drug (respt. Composit adl) clustering. The 3 by 3 heat map gives the distribution of patient in the intersection of cluster. The box first raw, first column corresponds to the number of patients that belongs to the first cluster for the drug dose and for first cluster for the adl score. The box first raw, second column corresponds to the number of patients that belongs to the first cluster for the drug dose and for the second cluster for the adl score, and so on.

### Nonlinear Dynamic of DDB & Score coupling with MGTX

To test the response of the patient score to a DDB, we computed the mean and standard deviation of three characteristics of the DDB per cluster: (i) the duration of the DDB in months – see Figure 10 right panel, (ii) the amplitude of the DDB in mg see Figure 11 left panel and (iii) the corresponding decay of score points - see Figure 11 right panel.

**Figure 10.**
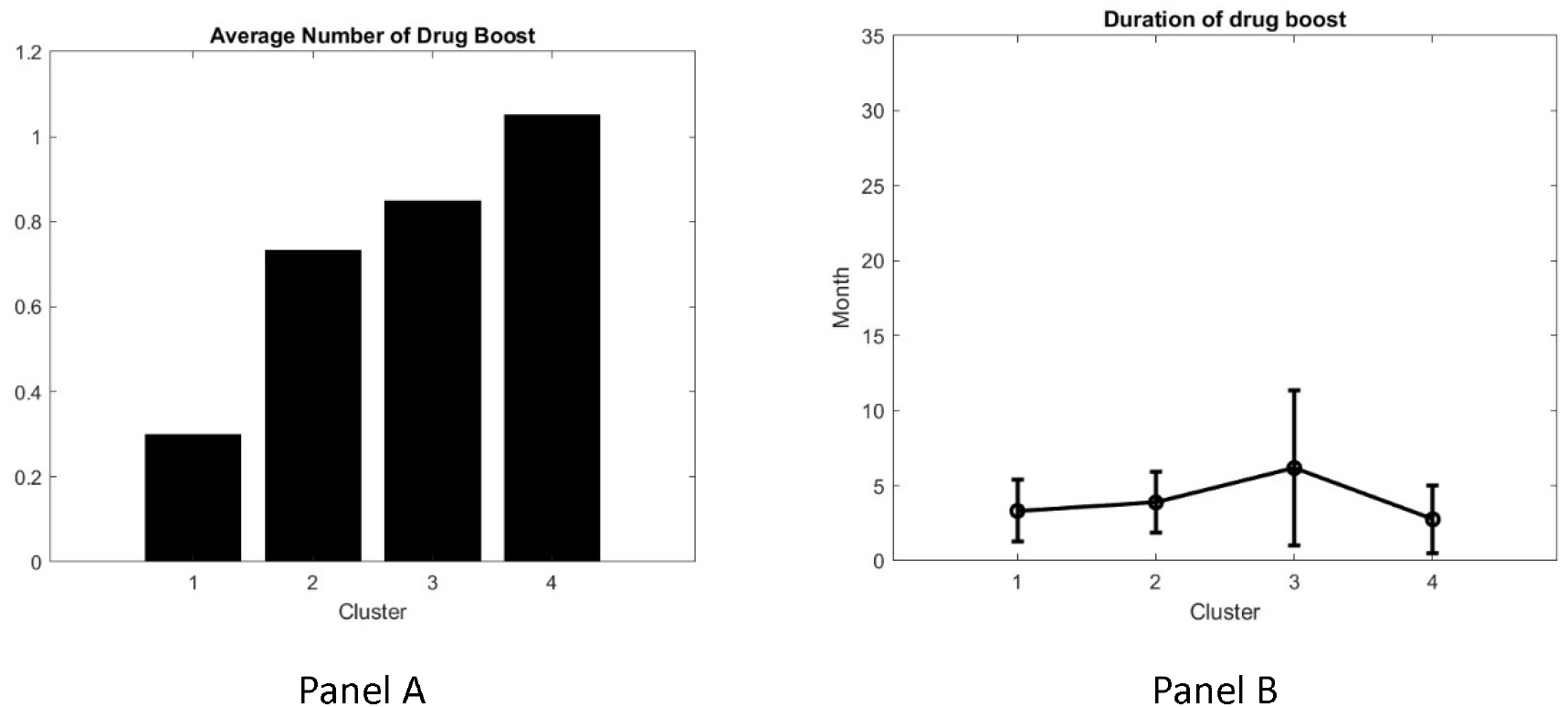
Average number of DDB of patient daily drug per cluster (panel A and corresponding average duration of the DDB per cluster (panel B)

**Figure 11.**
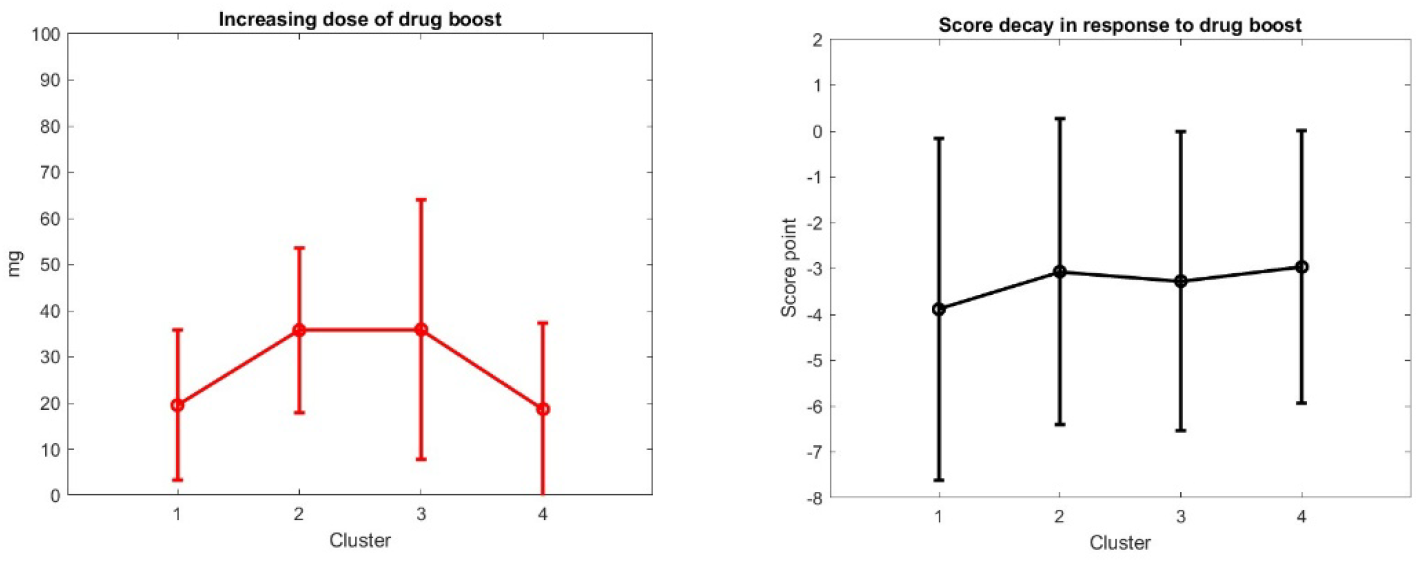
Average increase of drug dosage of a DDB in mg per patient per cluster (panel A) and average response of the score decay per DDB per cluster (panel B)

The average number of DDB per patient per cluster is as expected low for cluster one, see Figure 10 A. The results for the DDB characteristics corresponding to cluster 4 are particularly striking. The drug dosage is close to saturation and the opportunity to increase the drug dosage is limited. Clusters 2 and 3 are characterized by an increasing duration of DDB, a larger increase of the DDB and a lower response of the score decay. However, these results are sensitive to the smaller set of subjects in cluster 2 and 3 with large standard deviations and do not allow a robust mapping of the DDB to Delta Score. For patients in this category a larger data set would be required.

## 4. Discussion

The present study introduces a novel framework for clinical trial outcome prediction by leveraging patient trajectories—a method that transforms longitudinal clinical data into dynamic, individualized response maps. By integrating a unified clustering approach with a rigorously defined predictability index, our methodology not only distinguishes differences in treatment responses among MG patients within a clinical trial but also lays the groundwork for constructing digital twins. This innovative strategy captures the complex, time-dependent evolution of patient metrics specifically, QMG, MG-ADL scores, and daily prednisone dosages and provides a quantifiable means to assess treatment efficacy. The mathematical modeling, dynamic clustering, and personalized response mapping offers a powerful alternative to traditional, static approaches, which could be applied to the design and analysis of clinical trials, most importantly in rare diseases.

Our approach demonstrated that patient outcomes could be effectively stratified into distinct clusters that correlate with key clinical features such as age, smoking history, and thymectomy status. These clusters not only mirror known clinical relationships—for instance, the beneficial impact of thymectomy—but also reveal nuanced patterns of drug dosage response that traditional methods overlook, the variation in time of response to prednisone escalation among subjects, which has not been evaluated in clinical trials of MG patients. The establishment of a predictability index further enables the evaluation of treatment consistency over time, addressing one of the critical challenges in clinical trial modeling: the variability inherent in small, heterogeneous patient populations.

Despite these promising findings, the study acknowledges several limitations. The model’s predictive utility is constrained by the inherent noise of clinical data, including inconsistencies in data acquisition and protocol-driven variability in treatment adjustments. The drug dosage protocol relied on patient scores, acknowledging the moderate correlation between QMG and MG-ADL [18, 19, 20]. The adoption of Minimal Manifestation Status reflects the field’s recognition that optimal patient status does not necessarily equate to zero QMG and MG-ADL scores. Notably, MG-ADL exhibits a “floor effect,” limiting its ability to differentiate between patients with low disease severity, particularly within a clinical trial context. For example, patients with minimal clinical findings (e.g., slight weakness on arm extension) may exhibit normal functional capacity, leading to discrepancies between QMG and perceived well-being. However, we believe that there are ways to improve the accuracy of clinical scores using video acquisition and artificial intelligence [21-25]. There is inherent noise within the dataset, which limits the achievable precision of probabilistic estimates for factors, such as age, BMI, and smoking, on drug responsiveness.

The concept of trajectory may inspire several techniques to proceed with modeling. It is relatively straightforward to model the response of the score to the drug dosage using a simple logistic model or sigmoid curve: see Figure A5

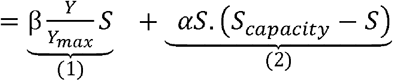

(1) and (2) may operate at different time scales: α is the time scaling factor that drives how fast the score may deteriorate without drug. is the time scaling factor that drives the delay to observe the drug effect on score.

The potential equilibrium and end point of the sigmoid equation is

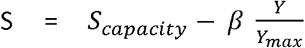

The sigmoid curve is by no means the representation of the response of the autoimmune system to the daily drug. As a matter of fact all assumptions on the behavior of the autoimmune response, neuromuscular transmission defect, and biological effect of drug sum up to the decision tree algorithm that defines the function Y(S,t) that is an observation of what the treatment protocol does for the patient. The sigmoid curve is simply a surrogate that enforces the saturation effect of drug and factor an indicator of the disease severity expressed as *S*_*capacity*_

We observed that this phenomenological model exhibits limited predictive utility due to several factors. Firstly, the immune system response is inherently complex and subject to change over the nearly three-year duration of the treatment trial. Secondly, the model lacks true predictive power without a more nuanced understanding of how drug dosage boosts (DDBs), administered in response to sudden disease worsening reflected by QMG and ADL score increases.

To address this, we shifted our focus towards a localized analysis, isolating DDBs for each patient and quantifying the corresponding score responses from the dataset. We hypothesize that a sufficiently large database of these localized response maps will provide the fundamental building blocks (“basic atoms”) necessary to reconstruct individual patient responses to daily drug dosage throughout the treatment period.

While we have not yet achieved comprehensive model construction due to data limitations, nor have we optimized score input by identifying the optimal combination of MG-ADL and QMG scores for treatment guidance, we remain optimistic that the aggregation of data from multiple clinical trials will enable the development of a robust predictive model.

Although our modeling does not proport to reflect the autoimmune pathology of MG, we do think there is the potential to reveal potential biological insights. Modeling of normal and autoimmune pathology is an active line of investigation [26,27, 28]. We appreciated a distinct pattern of treatment-response in a cohort of patients. Although variations in clinical response to corticosteroids are well appreciated for MG [29] and other autoimmune and inflammatory diseases [30], we found that higher doses overcome some level of poor response, while some patients reach a plateau of improvement regardless of drug dose. This suggests that drug metabolism relates to treatment response, which is consistent with findings of metabolomic analysis of MGTX subjects that indicated xenobiotic pathways were related to poor outcomes.

## Conclusion

In conclusion, this study paves the way for a paradigm shift in clinical trial design for myasthenia gravis and similar conditions. By capturing the dynamic interplay between patient characteristics and treatment responses, our trajectory-based approach not only enhances the precision of outcome predictions but also informs the development of personalized therapeutic strategies.

Furthermore, while our trajectory-based clustering provides an innovative lens through which to view patient responses, its full potential hinges on the availability of larger, more diverse datasets that can refine the model’s parameters and enhance its generalizability. As data integration and computational power continue to evolve, this method holds significant promise for improving patient care and advancing precision medicine in the realm of MG but easily can be generalized to many other disorders. Future work will focus on integrating additional data sources—such as real-time digital biomarkers, video-based assessments, and electronic medical records—to further validate and optimize our modeling approach to lead to a digital twin.

## Data Availability

All data produced in the present study are available upon reasonable request to the authors.

## Acknowledgements

The work was supported in part by the MGNet a member of the Rare Disease Clinical Research Network Consortium (RDCRN) NIH U54 NS115054. Funding support for the DMCC is provided by the National Center for Advancing Translational Sciences (NCATS) and the National Institute of Neurological Disorders and Stroke (NINDS). Additional support was provided by NSF-I Corp award 838792 (47354/1/CCLS 91906F). Dr. Garbey is CEO of Care Constitution, which supported digital tools and AI algorithms.

## Appendix

**Figure A1.**
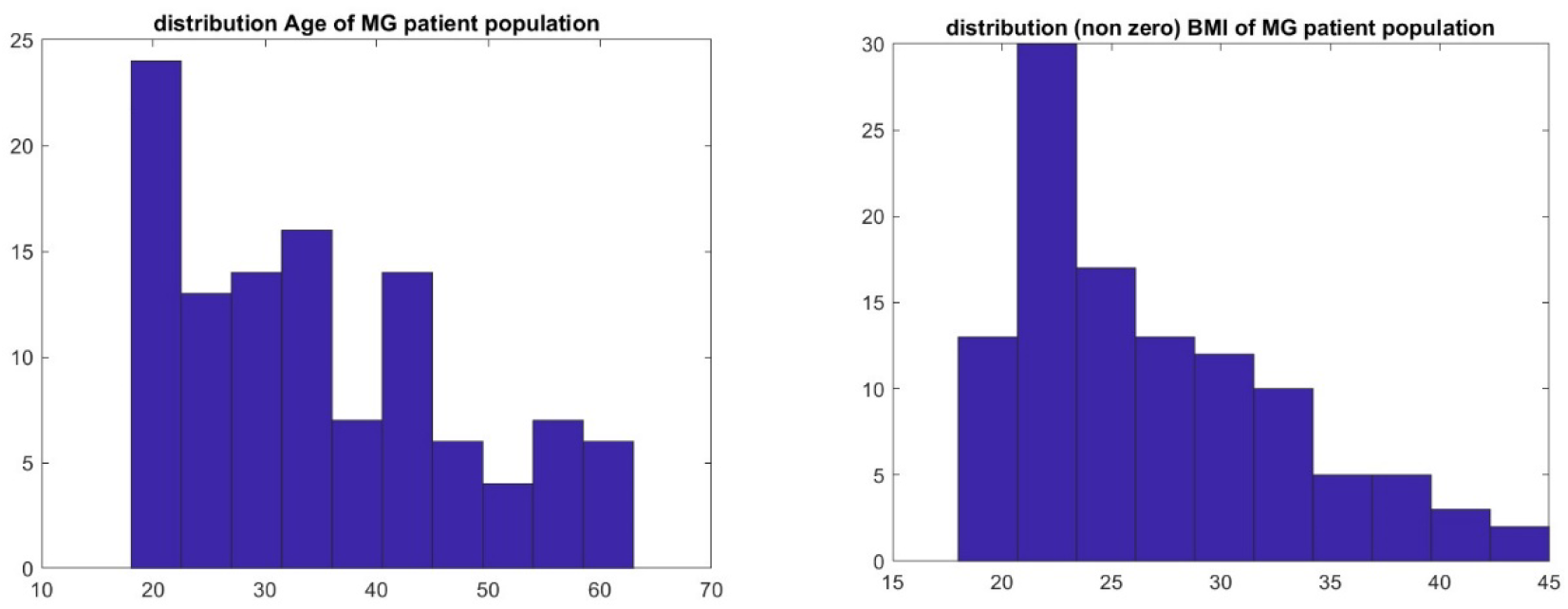
Properties of the MGTX patient population with respect to Age (Panel A) and BMI (Panel B).

**Figure A2.**
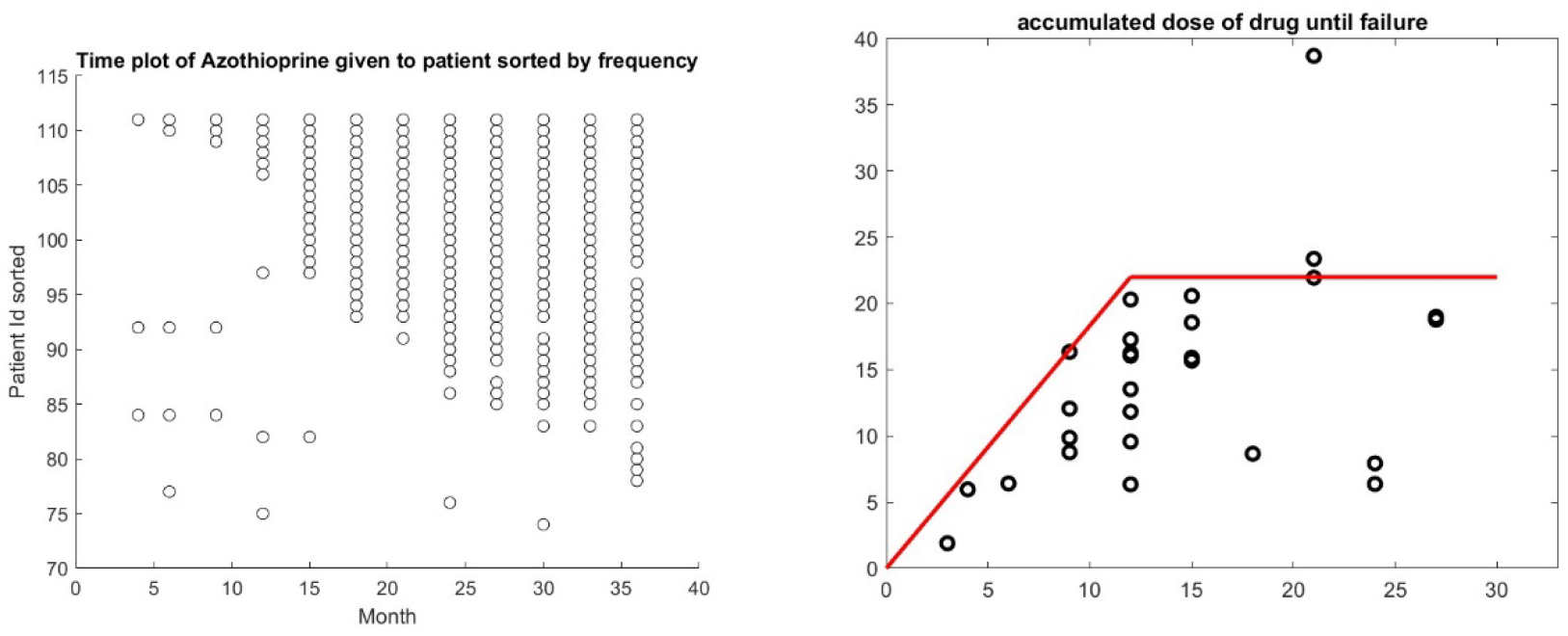
Temporal behavior of dose of azothiopine given to patient during the MGTX trial: once a patient is on this drug, he usually stays on it. Failure being define as the time patient starts to be on azothiopine, it seems that there is a saturation effect of the accumulated dose of prednisone that corresponds to the time of failure of the treatment.

**Figure A3.**
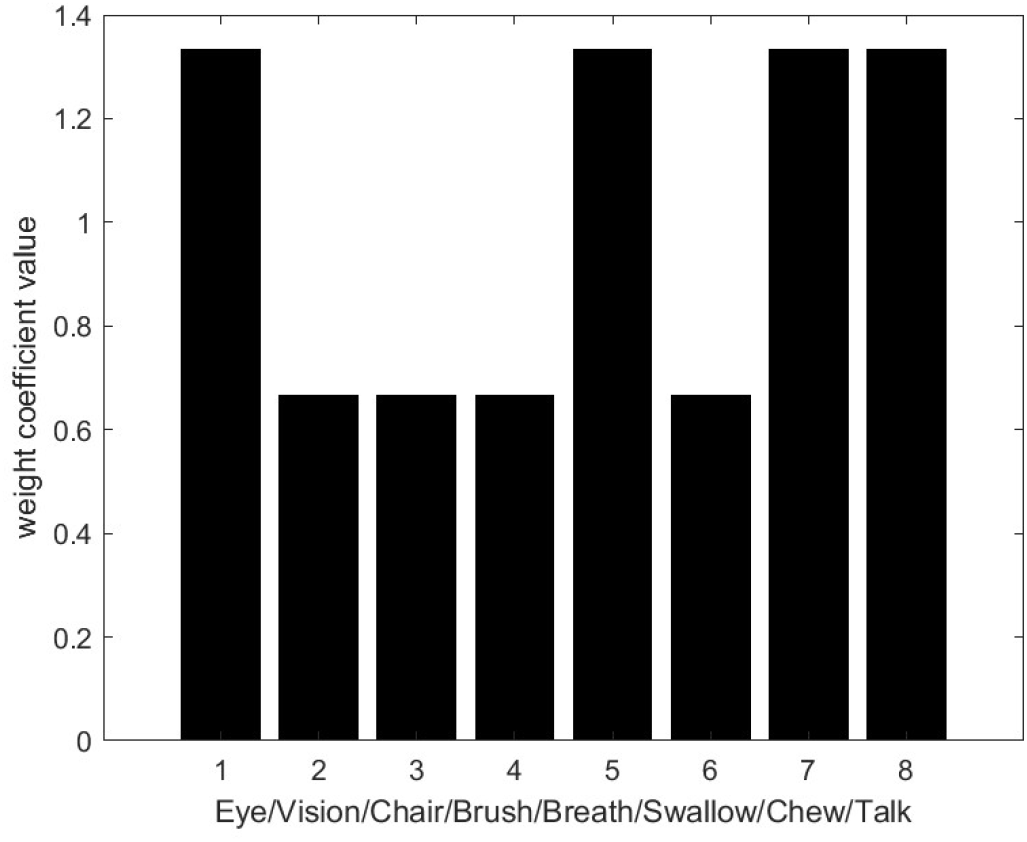
Optimum weight function for the composite ADL score.

**Figure A4.**
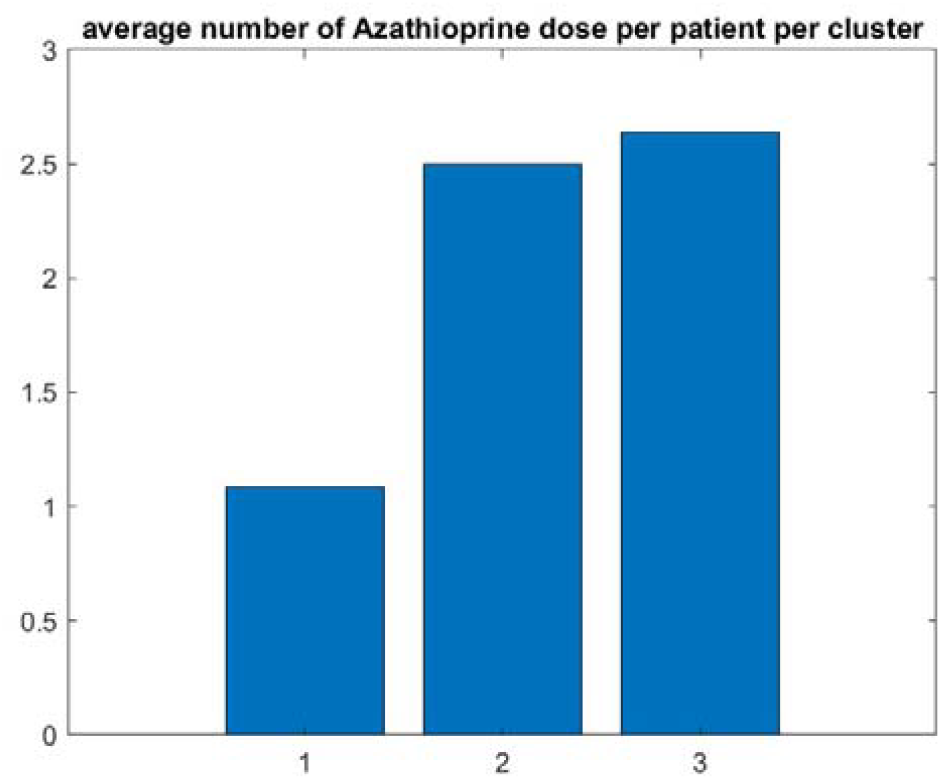
Requirement on Azathioprine with the 3 cluster trajectories for the ADL composite outcome is consistent with the result with the drug trajectory clusters in Figure 4 (MGTX Patients).

**Figure A5.**
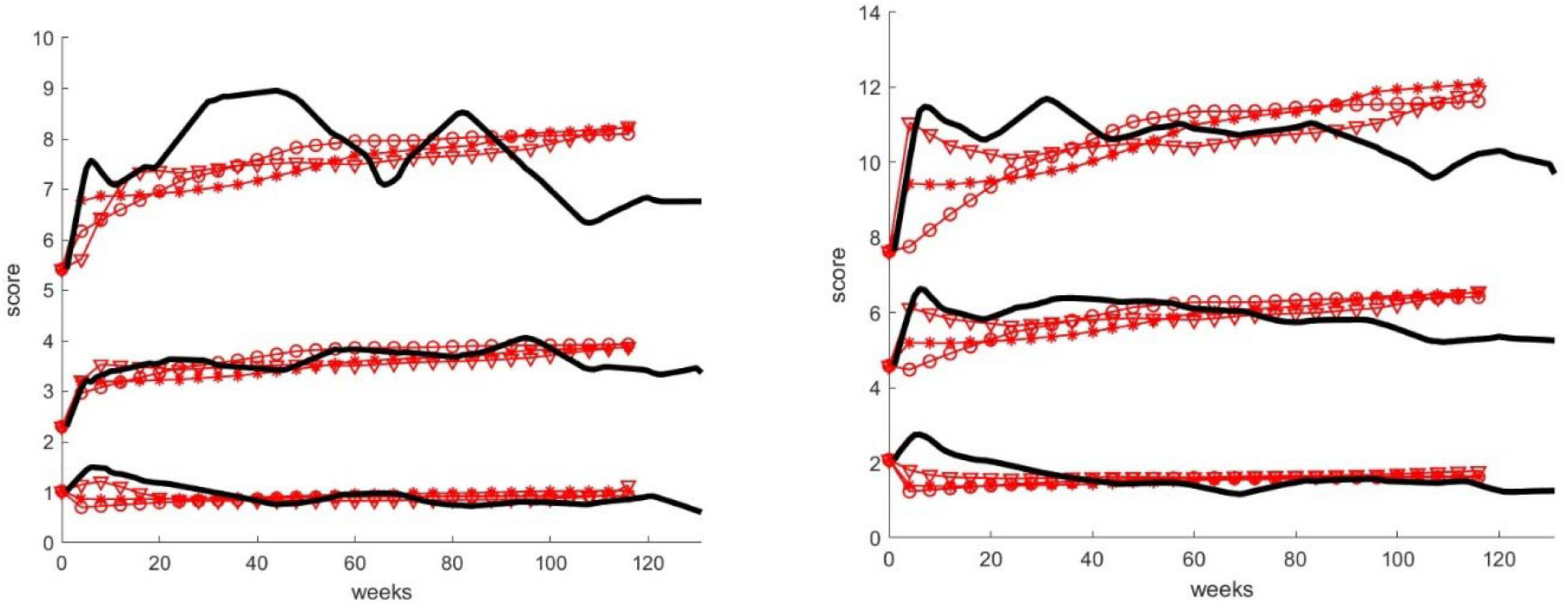
Model fitting for all three cluster scores curves. O, *, v are respectively for drug cluster input 1 to 3.

**Table 1:** best “growth rate” α and the “carrying capacity” *S*_*capacity*_ to match the score curves corresponding to each pair of cluster intersections (Si,Yj), i=1..3, j=1..3.

| $S_i \setminus Y_j$ | 1 | 2 | 3 |
| --- | --- | --- | --- |
| 1 | (2.1 , 0.96) | (0.5 , 4.1) | (0.25 , 8.4) |
| 2 | (2 , 1.2 ) | (0.5 , 4.6) | (0.25 , 9.6) |
| 3 | (2.4 , 1.2) | (0.5 , 5.3) | (0.25 , 11) |

